# Everyday functioning in a community-based volunteer population: Differences between participant- and study partner-report

**DOI:** 10.1101/2021.11.01.21265650

**Authors:** M. Verrijp, M.A. Dubbelman, L.N.C. Visser, R.J. Jutten, E.W. Nijhuis, M.D. Zwan, H.P.J. van Hout, P. Scheltens, W.M. van der Flier, S.A.M. Sikkes

## Abstract

**INTRODUCTION:** Impaired awareness in dementia due to Alzheimer’s disease and related disorders, made study partner-report the preferred method of measuring interference in ‘instrumental activities of daily living’ (IADL). However, with a shifting focus towards earlier disease stages and prevention, the question arises whether self-report might be equally or even more appropriate. This study aims to investigate how participant and study partner report IADL perform in a community-based volunteer population without dementia, and which factors relate to differences between participant and study partner report.

**METHODS:** Participants (N=3288; 18–97 years, 70.4% females) and their study partners (N=1213; 18–88 years, 45.8% females) were recruited from the Dutch brain research registry. IADL was measured using the Amsterdam IADL Questionnaire. Concordance between participant and study partner-reported IADL difficulties was examined using intraclass correlation coefficient (ICC). Multinomial logistic regressions were used to investigate which demographic, cognitive and psychosocial factors related to participant and study partner differences, by looking at the over- and underreport of IADL difficulties by the participant, relative to their study partner.

**RESULTS:** The vast majority of A-IADL-Q scores represented no difficulties for both participants (87.9%) and study partners (89.4%). Concordance between participants and study partners was moderate (ICC=.55, 95%CI=[.51, .59]), 24.5% (N=297) of participants overreported their IADL difficulties compared to study partners, and 17.8% (N=216) underreported difficulties. The presence of depressive symptoms (odds ratio (OR)=1.31, 95%CI=[1.12, 1.54]), as well as memory complaints (OR=2.45, 95%CI=[1.80, 3.34]), increased the odds of participants overreporting their IADL difficulties. Higher IADL ratings decreased the odds of participant underreport (OR=0.71, 95%CI=[0.67, 0.74]).

**CONCLUSION:** In this sample of community-based volunteers, the majority of participants and study partners reported no major IADL difficulties. Differences between participant and study partner were, however, quite prevalent, with subjective factors indicative of increased report of IADL difficulties by the participant in particular. These findings suggest that self- and study partner-report measures may not be interchangeable, and that the level of awareness needs to be taken into account, even in cognitively healthy individuals.

## Introduction

Decline in cognitively complex ‘instrumental activities of daily living’ (IADL) is among the first observable indicators of cognitive problems in everyday life, and is considered inherently clinically meaningful. Previous studies have shown that study partners report a decline in IADL in preclinical Alzheimer’s disease (AD), even before cognitive problems can be detected by standard cognitive testing (1-3). IADL instruments are considered an important outcome in clinical trials and intervention studies, even more so as the field shifts towards preclinical phases of AD and related disorders (4).

Due to impairments in awareness in persons with dementia (5), (I)ADL functioning has traditionally been assessed using study partner-report questionnaires (6-14). However, it has been suggested that study partner-report may be influenced by study partner depression, anxiety and caregiver burden (15-17). With a shift towards studying cognitively normal or “at risk” individuals, one might assume that participants are able to reliably reflect on their own level of functioning, as they are thought to have accurate or potentially heightened awareness of their functional and cognitive abilities (5, 18). In such populations, participant-report may therefore be a more appropriate and direct assessment method in cognitively normal individuals (12, 15, 17).

The validity of participant-reported IADL in preclinical or “at risk” populations is not yet clear. Alterations in awareness may already occur in preclinical populations and may lead to discordance between participants and study partners, where one may report more or less impairments than the other. Only few studies compared participant with study partner-reported functioning in community-based populations (11, 19). Studies in patients with dementia or MCI seem to indicate that there is a greater chance of discordance when participants have depressive symptoms or lower objective cognition (12, 19-22), and that discordance decreased when participant and study partner are living together (16). Studies investigating the interplay of these factors in cognitively normal populations are scarce. Furthermore, findings are difficult to compare between studies, due to differences in IADL measurements and in the definition and operationalization of concordance and discordance.

The Amsterdam IADL Questionnaire (A-IADL-Q) was developed as a study partner-rated questionnaire, and has been extensively validated in memory clinic and international aging populations (20, 23-30). It is not yet known how the participant-report version of the A-IADL-Q performs, and how it relates to study-partner report. The aim of this study is to investigate how the participant- and study partner-reported versions of the A-IADL-Q perform in a community-based population, without dementia. Second, we aim to investigate what factors relate to differences between participant- and study partner-reported IADL functioning.

## Materials and Methods

### Participant selection and study design

Participants were recruited through the Dutch online brain research registry (Hersenonderzoek.nl), which is an online platform for people interested in cognition and brain-related research (31). All eligible registrants were invited by e-mail to participate in the study. The only inclusion criterion was being 18 years or older. Those who self-reported to have received a dementia-related diagnosis (i.e., dementia or mild cognitive impairment [MCI]) were excluded.

Data collection started in August 2018 and ended in December 2018. The study was approved by the medical ethical committee of the VU University Medical Center. The participants provided consent via Hersenonderzoek.nl. Since study partners were not recruited through Hersenonderzoek.nl, they provided consent prior to completing the online IADL questionnaire.

### Measures

#### Amsterdam IADL Questionnaire

The main outcome measure was the Amsterdam IADL Questionnaire (A-IADL-Q). The A-IADL-Q was developed as a study partner-report instrument aimed at measuring problems in cognitively complex everyday functioning (23). For the current study, we adapted the study partner-report version to a participant-report version. Both versions consist of the same 30 items, covering a broad range of cognitive IADLs. Each item assesses difficulty performing an activity due to cognitive problems, such as memory, attention, or executive functioning. Item responses were rated on a five-point Likert scale, ranging from “no difficulty in performing this activity” (0) to “no longer able to perform this activity” (4). The total score is calculated using item response theory (IRT), assuming a single underlying construct (32), that is, IADL functioning, ranging from disability to ability. Total scores range from 20 to 70, and were reversed so that higher scores reflect better IADL functioning. A cutoff for dementia was previously placed at 51.4 (24), while scores above 60 were considered to indicate no IADL difficulties (33). The study partner-report version of the A-IADL-Q has undergone extensive validation, showing good content and construct validity, high internal consistency, high test-retest reliability, good responsiveness to change and able to measure IADL across cultures and languages (20, 24-26, 29). The study partner version of the A-IADL-Q also includes questions about the type of relation to the participant and cohabitation. Study partners were classified as spouses, children, siblings or ‘other’. Study partners in the ‘other’ category included friends, coworkers, or other family members.

#### Other measures

Cognitive functioning was assessed using the Cognitive Online Self-Test Amsterdam (COST-A), an online cognitive self-test developed and validated by Van Mierlo et al. (34). The COST-A included 10 tasks, covering multiple cognitive domains. Performance on each of the 10 tasks was standardized into a Z-score, where higher scores indicate better cognition. These Z-scores were averaged into a global score representing overall cognitive functioning. Visser et al. (2021) provides a more detailed description of the COST-A.

Additionally, a single yes/no question (“Do you have memory complaints?”) assessed subjective memory complaints. Depressive symptoms were assessed with the five-item short form of the Geriatric Depression Scale (GDS5) (35). with higher scores indicating more depressive symptoms. Education level was classified as low–medium (up to high school) and high education (college degree).

### Defining awareness of IADL functioning

In line with other studies, we defined concordance based on the discrepancy between participant- and study partner-report (5). We categorized concordance into three groups, based on a previously determined clinically meaningful difference of 2.4 points (36): (1) concordance between dyads; (2) discordance between dyads with the participant ‘over reporting’ difficulties (i.e., scoring ≥2.4 points lower than their study partner); and (3) discordance between dyads with the participant ‘underreporting’ difficulties (i.e., scoring ≥2.4 points higher than their study partner).

### Statistical analyses

Demographic differences between study partners and participants were tested using independent t-tests or chi-square tests. The frequency of IADL difficulties among cognitively normal participants and their study partners was determined. Then, in separate linear regression analyses, A-IADL-Q scores of both raters were associated with age, education, objective cognitive functioning, subjective cognitive functioning, and depressive symptoms.

The intraclass correlation coefficient (ICC) was computed to examine concordance between participants and study partner ratings. According to the criteria of Koo et al., an ICC <.5 shows poor concordance, an ICC of .5–.75 shows moderate, and >.75 shows good concordance (37).

Using stepwise multinomial logistic regression models with backward selection, we investigated which factors related to concordance and discordance between dyads. The included variables were participants’ education level, sex, age, COST-A scores, memory complaints, and GDS5 total score, study partner-reported IADL functioning, the type of relationship, cohabitation (yes/no), and the absolute age difference between dyads. For this analysis, COST-A scores were dichotomized into normal (>-1.5SD) and low (≤-1.5SD) cognitive functioning. All analyses were performed in R version 4.0.3 (38).

## Results

Of the 11,060 eligible registrants, 4,817 individuals (44%) were interested in participation and received study instructions. After receiving instructions, 3,288 (68%) completed the participant-reported A-IADL-Q. On average, participants were 61.0±12.1 years old and the majority were female (2,315; 70.4%). Approximately half the participants experienced memory complaints. Table 1 displays all participant and study partner characteristics.

**Table 1.** Participant and study partner characteristics

|  | Participants<br>(N = 3,288) | Dyads<br>(N = 1,213) |  |
| --- | --- | --- | --- |
|  |  | Participants | Study partners |
| Age, mean (SD) | 61.0 (12.1) | 62.5 (11.1) | 58.8 (14.2) |
| Range | 18–97 | 18–93 | 18–88 |
| Female, n (%) | 2,315 (70.4) | 828 (68.3) | 556 (45.8) |
| High level of education, n (%) | 2,323 (70.7) | 854 (70.4) | — |
| A-IADL-Q score, mean (SD) | 65.9 (4.8) | 65.9 (4.7) | 66.1 (4.6) |
| Range | 40.9–70.0 | 40.7–70.0 | 42.7–70.0 |
| Memory complaints present,<sup>1</sup> n (%) | 1,429 (47.5) | 586 (49.9) | — |
| COST-A,<sup>2</sup> abnormal performance ($\leq -1.5SD$), n (%) | 225 (7.6) | 86 (7.5) | — |
| GDS5,<sup>1</sup> median (IQR) | 0 (0–1) | 0 (0–1) | — |
| Type of relationship, n (%) |  |  |  |
| Spouse |  | 956 (78.8) |  |
| Child |  | 155 (12.8) |  |
| Sibling |  | 32 (2.6) |  |
| Other |  | 70 (5.8) |  |
| Duration relationship, n (%) |  |  |  |
| <5 years |  | 33 (2.7) |  |
| 5–10 years |  | 58 (4.8) |  |
| >10 years |  | 1,119 (92.5) |  |
| Living together, n (%) |  | 960 (79.3) |  |

‘—’ denotes that data was not available. <sup>1</sup> Data available for 3,011 participants, 1,175 of whom were part of a dyad. <sup>2</sup> Data available for 2,945 participants, 1,149 of whom were part of a dyad.
*Abbreviations: A-IADL-Q, Amsterdam Instrumental Activities of Daily Living Questionnaire; COST-A, Cognitive Self-Test Amsterdam; GDS5, 5-item Geriatric Depression Scale; IQR, interquartile range; SD, standard deviation.*

For 1,213 participants (36.9% of complete sample), the A-IADL-Q was also completed by a study-partner (participant and study partner pairs will be referred to as ‘dyads’). Participants who were part of a dyad were older (*p* < .001) and were more often male (*p* = .046) than those who were not. Within dyads, the participants were older (*p* < .001) and more likely to be female (*p* < .001), compared to study partners.

### IADL difficulties in a cognitively normal population

Figure 1 shows the distribution among dyads of participant- and study partner-reported A-IADL-Q scores. The participant-reported A-IADL-Q scores (65.9±4.8) did not differ from the study partner-reported A-IADL-Q scores (66.1± 4.6; *p* = .186). Virtually all participants (3,232/3,288; 98.3%) and study partners (1,195/1,213; 98.5%) reported A-IADL-Q scores above a previously established cutoff for dementia (total score of 51.4). Moreover, the vast majority of both participant-reported (87.9%) and study partner-reported (89.4%) total scores where higher than 60, indicating no difficulties.

**Figure 1.**
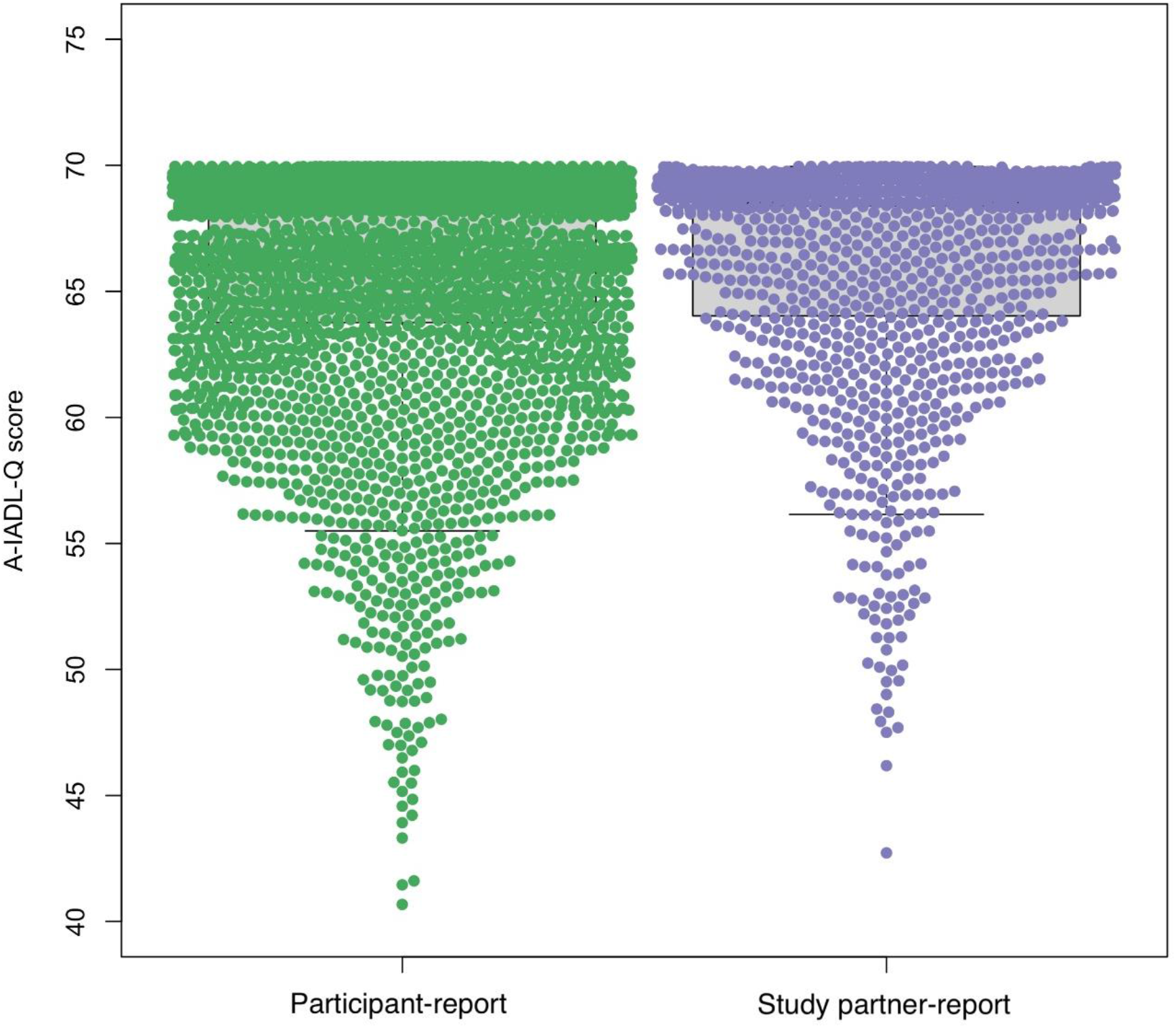
A-IADL-Q total score distribution among participants (N=3,288, in green) and study partners (N=1,213, in purple).

**Figure 2.**
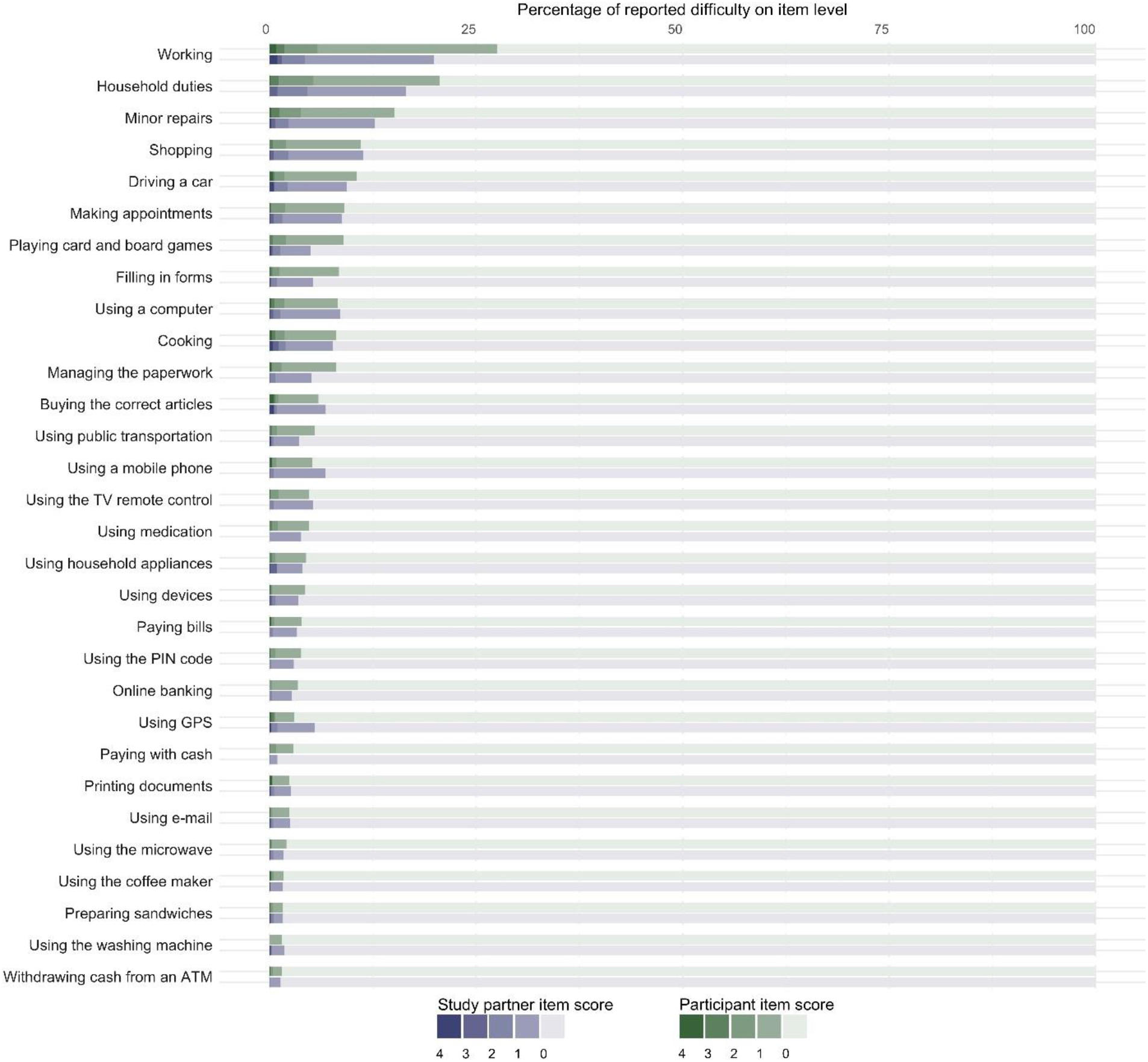
Stacked bar chart showing the percentage of participants (in shades of green) and study partners (in shades of purple) who reported difficulties (N=1,213). The dark shades represent difficulty with the activity: “no longer able to perform this activity” (4), “much more difficulty” (3), “more difficulty” (2), and “slightly more difficulty” (1). The lightest shade represents “no difficulty in performing this activity” (0).

Next, we looked at IADL difficulties at item level. Half of all participants (1,750/3,288, 53.2%) and study partners (722/1,213, 59.5%) reported no difficulties in any activity. Those who did report difficulties, mostly did so in only one activity (35.2% of participants, 35.8% of study partners). **Error! Reference source not found**. shows the percentage of participants and study partners who reported difficulties for each IADL activity. Most frequently reported IADL difficulties for both participants and study partners were working (26.9% and 19.9%, respectively), household duties (22.2% and 16.5%, respectively) and making minor repairs at home (16.4% and 12.7%, respectively).

Table 2 shows the associations between age, education level, cognitive complaints, COST-A, GDS and participant- and study partner-reported IADL performance. Higher age was associated with lower A-IADL-Q scores, and higher education with better A-IADL-Q scores, but associations were weak. For example, with every 10 years increase in age, A-IADL-Q participant- and study partner-reported scores decreased with 1.2 and 1.8 points, respectively. Both participant- and study partner-reported A-IADL-Q scores were more highly associated with COST-A scores, memory complaints, and GDS. Higher COST-A scores, indicating better cognitive functioning, were associated with better IADL functioning, whereas a higher GDS, indicating more depressive symptoms, and presence of memory complaints were associated with worse IADL functioning. Associations with age, education and COST-A scores were comparable for participant- and study partner-report, whereas associations with GDS and memory complaints were more strongly associated with participant-reported IADL scores.

**Table 2.**
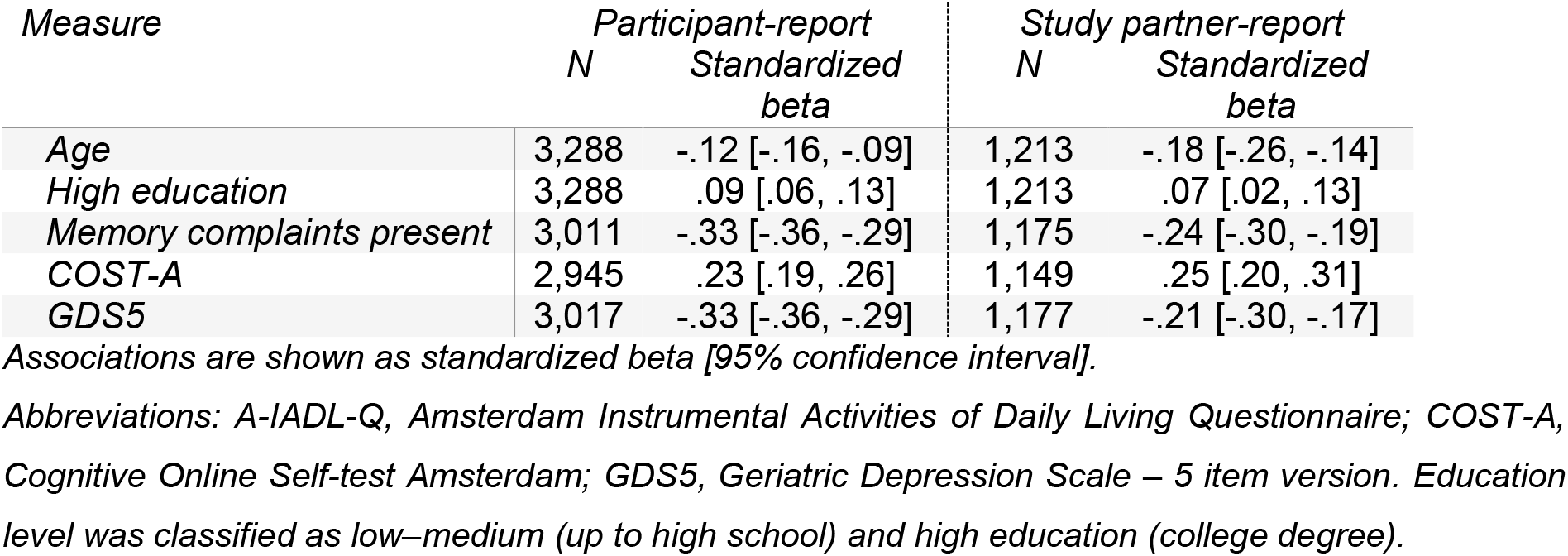
Linear regressions to investigate associations with participant- and study partner-reported IADL performance

**Table 3.** Multivariable multinomial logistic regression models comparing study partners reporting more IADL difficulties than participant (N=216) and participant reporting more IADL difficulties than study partner (N=297), compared to agreement between participant and study partner (N=700).

| Predictor | Study partner > Participant<br>(N=216) |  | Participant > study partner<br>(N=297) |  |
| --- | --- | --- | --- | --- |
|  | OR [95%CI] | P | OR [95%CI] | P |
| COST-A ≤-1.5 SD | 0.47 [0.21, 1.07] | .070 | 1.36 [0.78, 2.39] | .283 |
| A-IADL-Q (study partner-report) | 0.71 [0.67, 0.74] | < .001 | 1.04 [0.99, 1.09] | .148 |
| Memory complaints present | 0.76 [0.50, 1.15] | .194 | 2.44 [1.80, 3.32] | < .001 |
| High education | 0.92 [0.60, 1.40] | .689 | 1.30 [0.93, 1.80] | .121 |
| Absolute age difference between dyads<br>in years | 1.00 [0.97, 1.04] | .924 | 1.01 [0.98, 1.04] | .924 |
| Age in years (participant) | 1.01 [0.99, 1.03] | .467 | 1.01 [0.99, 1.02] | .272 |
| Female sex (participant) | 0.74 [0.53, 1.02] | .159 | 1.08 [0.78, 1.49] | .661 |
| GDS5* | 0.58 [0.50, 0.68] | < .001 | 1.31 [1.12, 1.53] | < .001 |
| Type of relationship, study partner is a:† | 2.19 [0.63, 7.60] | .216 | 0.83 [0.30, 2.27] | .716 |
| Child | 0.75 [0.13, 4.35] | .744 | 0.57 [0.18, 1.85] | .350 |
| Sibling | 0.81 [0.22, 2.98] | .755 | 0.63 [0.24, 1.68] | .355 |
| Other |  |  |  |  |
| Dyads live together | 1.58 [0.70, 3.57] | .277 | 1.04 [0.57, 1.90] | .898 |

*Abbreviations: COST-A, Cognitive Online Self-Test Amsterdam; GDS5, 5-item short form of Geriatric Depression Scale; OR, odds ratio; 95%CI, 95% confidence interval. Concordance was used as a reference group (N=700). \* more depressive symptoms; † Using spouse as a reference category.*

### Concordance and discordance between dyads

There was a moderate consistency between participant- and study partner-reported IADL functioning (intraclass correlation coefficient = .55, 95%CI = [.51, .59], *p* < .001; see Supplementary Figure 1). Of all 1,213 dyads, 700 (57.7%) were in concordance. Two hundred sixteen participants (17.8%) underreported difficulties, compared to their study partners, and 297 participants (24.5%) over reported IADL difficulties, compared to their study partners. Compared to concordant dyads, participants with memory complaints (odds ratio (OR) = 2.44, 95%CI = [1.80, 3.32], *p* < .001) and with a higher GDS (OR = 1.31, 95%CI = [1.12, 1.53], *p* = .001) were more likely to overreport IADL difficulties. Participant underreport was less likely when there were fewer IADL difficulties (OR = 0.71, 95%CI = [0.67, 0.74], *p* < .001). Thus, concordance was more likely when the participant did not experience memory complaints, had lower GDS scores and when IADL performance was higher. Participant’s education, age, gender and COST-A scores were not related to concordance between dyads.

## Discussion

In this study, we showed that the majority of IADL scores fell within the range of normal IADL functioning in this community-based population, but that discordance among dyads was quite prevalent. A small subgroup reported subtle IADL difficulties, which was associated with participants’ older age, lower education, worse cognitive performance, presence of memory complaints, and more depressive symptoms, for both participant and study partner-report. Moderate concordance between participant and study partner-reported IADL was found with discordance being more likely when the participant experienced memory complaints, depressive symptoms, and lower IADL performance.

Whilst the large majority of participant and study partner-reported IADL functioning fell within the range of normal IADL functioning, approximately a tenth of both participants and study partners scored below the previously established cutoff for normal IADL functioning (33). This prevalence of impaired IADL is comparable to other population-based studies (21, 39-41). For example, Scheel-Hincke and colleagues (40) reported a prevalence of impaired IADL of 12 to 20% in Western Europe, with impaired IADL defined as presence of any difficulties. Another population-based study by Pudaric and colleagues (41) reported a prevalence of impaired IADL (inability to carry out shopping, cooking or housework) of 6 to 11%. Despite this comparable prevalence of abnormal IADL functioning, it is important to note that approximately half of our population reported more subtle difficulties. If we applied the definition of Scheel-Hincke et al. (40), the prevalence of impaired IADL in our study would be approximately 50%, which is substantially higher than the prevalence that they reported. There are two potential explanations for this difference: first, we included more activities, and, second and more importantly, we included more cognitively complex activities than other studies. This is illustrated by the fact that most problems were reported in working, household duties and making repairs, which are especially cognitively complex (26). These activities were not reported for other IADL scales. For example, a population-based study which assessed five IADL items (42), reported most problems for shopping. In our population, problems with shopping was fourth most prevalent. We found a higher proportion of difficulties for more complex activities, supporting the notion that including more complex activities enabled detection of more fine-grained difficulties in IADL functioning.

With regard to potential sources of bias in the report of IADL functioning, we found low associations between both study partner and participant-reported IADL functioning and age and education, a finding which is in line with previous validation studies for the study partner version of the A-IADL-Q (20, 26, 29). Participant- and study partner-report were similarly associated with objective cognitive performance, but participant-report was more strongly related to depressive symptoms as well as subjective cognitive performance (i.e., presence of memory complaints). Consistent with recent literature suggesting that study partners are better able to assess the participants’ functioning than the participant themselves (43), our findings imply that study partner-report might be less biased than participant-report by participant-related subjective factors.

Our findings demonstrated only a moderate concordance between dyads. While the distributions of study partner- and participant-reported IADL scores were largely similar, we found a moderate ICC, and a high proportion of discordance (either over or underreport). Other studies have also shown discordance in cognitively normal participants, and specifically participant overreport (11, 19, 21, 44). For example, a study by Okonkwo and colleagues (19) showed slight discordance between participant and study partner-report of specific finance-related IADLs. The proportion of discordance that we found in our study is substantially higher, which is probably due to differences in IADL measures, definitions of concordance and population differences. As opposed to Okonkwo and colleagues (19), who calculated concordance based on an individual item, we determined concordance based on a more global measure of IADL with a wider range of activities. We calculated concordance based on a clinical meaningful difference in total scores. Another potential explanation may be that, even though we used a population-based sample, we did not screen for cognitive impairment. As such, it is possible that there were participants who had subtle cognitive impairment, but did not meet criteria for MCI or dementia. Thus, while the proportion of discordance is difficult to compare to other studies, the fact that other studies also reported discordance, suggests that participant and study partner report might not be interchangeable.

The potential limited interchangeability is further supported by our results which indicate that concordance is influenced by memory complaints and depressive symptoms. Participants with memory complaints reported more difficulties, compared to their study partners. Participant overreport of memory complaints has previously been described as a heightened awareness (5), which is thought to characterize early stages of Alzheimer’s disease and related disorders (5, 45, 46). Following this theory, a subgroup of our study sample may have a heightened functional awareness. This idea is further supported by our finding that a large proportion of our sample had memory complaints, which may indicate a heightened memory awareness. While no other studies have investigated the effect of subjective cognitive functioning on the concordance of functional impairment, several studies (11, 19, 21, 44, 47-49) related objective cognitive functioning to concordance. These studies show that patients with poorer global cognition are more likely to underreport IADL difficulties. We did not find an significant association between concordance and objective cognition within our healthy volunteer population. This could be due to the fact that our population is presumably cognitively healthy, and lowered awareness may not occur until later disease stages (5, 50). Although not significant, in this population, lower cognitive performance seems to be related to a reduced odds for participant underreport. This might suggest that the subtle cognitive problems of these individuals do not interfere with their disease insight, but rather, that they increase their awareness. Furthermore, participants with depressive symptoms were more likely to overreport, and less likely to underreport, IADL difficulties. This is in line with the idea that negative self-perception in patients with depressive symptoms causes exaggeration of deficits (51), as has also been shown by Okonkwo and colleagues (19), who reported that underestimation of financial abilities was related to higher depressive symptoms. Thus, memory complaints and depressive symptoms both influence the participant’s report of their IADL difficulties, and need to be taken into consideration when using participant-reported IADL measures.

The findings discussed above may have important implications for study design decisions and should be considered carefully when considering the use of a participant-reported IADL instrument. Although a concordance of 60% might seem low, the majority of both participant and study partner-reported difficulties fell within the category of ‘no difficulties’. This crude overlap indicates that participant-report IADL can be useful in cognitively normal populations in cross-sectional studies. However, when a deterioration of cognitive functioning and subsequently everyday functioning is to be expected, study partner-report might provide a more reliable indication of change in IADL functioning. The combination of participant- and study partner-report can be used to establish awareness, which is informative since it has been shown to predict future disease progression (52, 53) and greater discordance seems to be related to a greater risk of Alzheimer pathology (5, 47). Thus, participant self-report can be used in cognitively normal populations, but should ideally be supplemented by study partner-report for longitudinal studies.

Some limitations should be considered when interpreting our findings. For lack of an objective IADL measure, we cannot ascertain whether participants indeed overreport their difficulties, or whether participants actually have IADL difficulties that the study partner does not yet notice. In other words, a heightened participant awareness may also reflect lowered study partner awareness. This caveat notwithstanding, the absence of an association with objective cognitive functioning could indicate that participant overreport is more strongly influenced by subjective than objective factors. Further, as study-partner report is generally considered a gold standard in dementia research and clinical practice (54), we used it as such in the current study. Another limitation is the selective nature of the volunteer registry, which consists mostly of highly educated and highly motivated individuals. This may limit generalizability to the general population. We did not include factors such as caregiver burden, personality traits or more detailed information on the amount of contact between the participant and the study partner. Future studies should consider to assess these factors to obtain more detailed insight into the accuracy of assessments and possible biases. Furthermore, follow-up studies are needed to determine the pivot point until which the participant is still able to reliably evaluate their own level of daily functioning.

An important strength of this study is the large sample of cognitively healthy volunteers, representing a large range of ages, from early adulthood to late life. We included detailed information about the level of IADL difficulties from both self and study partner-report in a cognitively healthy population, providing valuable new insights into the occurrence of more subtle IADL difficulties. Another strength is our use of a clinically meaningful cutoff to distinguish concordance from discordance. For this reason, we believe that discordance actually represented an important, non-negligible difference in IADL report.

In conclusion, our findings show a moderate concordance between participants and study partners in reporting IADL difficulties, with subjective factors influencing the level of concordance. These findings suggest caution in using self- and study partner-report measures interchangeably, even in cognitively healthy community-based samples. Our results suggests that participant report might be more related to subjective factors and that study partner report is less associated with these factors, possibly reflecting differing perspectives.

## Data Availability

All data produced in the present study are available upon reasonable request to the authors

## Acknowledgments

We thank the participants and their study partners for their time and participation in our study. The participant recruitment was accomplished through Hersenonderzoek.nl, a Dutch online registry that facilitates participant recruitment for neuroscience studies (www.hersenonderzoek.nl). Hersenonderzoek.nl is funded by ZonMw-Memorabel (project no 73305095003), a project in the context of the Dutch Deltaplan Dementie, Gieskes-Strijbis Foundation, the Alzheimer’s Society in the Netherlands and Brain Foundation Netherlands. Funding for the data collection and research on which this manuscript is based was provided by the Stichting Stoffels-Hornstra.

## Supplementary Material

### Relationship between participant and study partner-reported IADL scores

**Figure 1.**
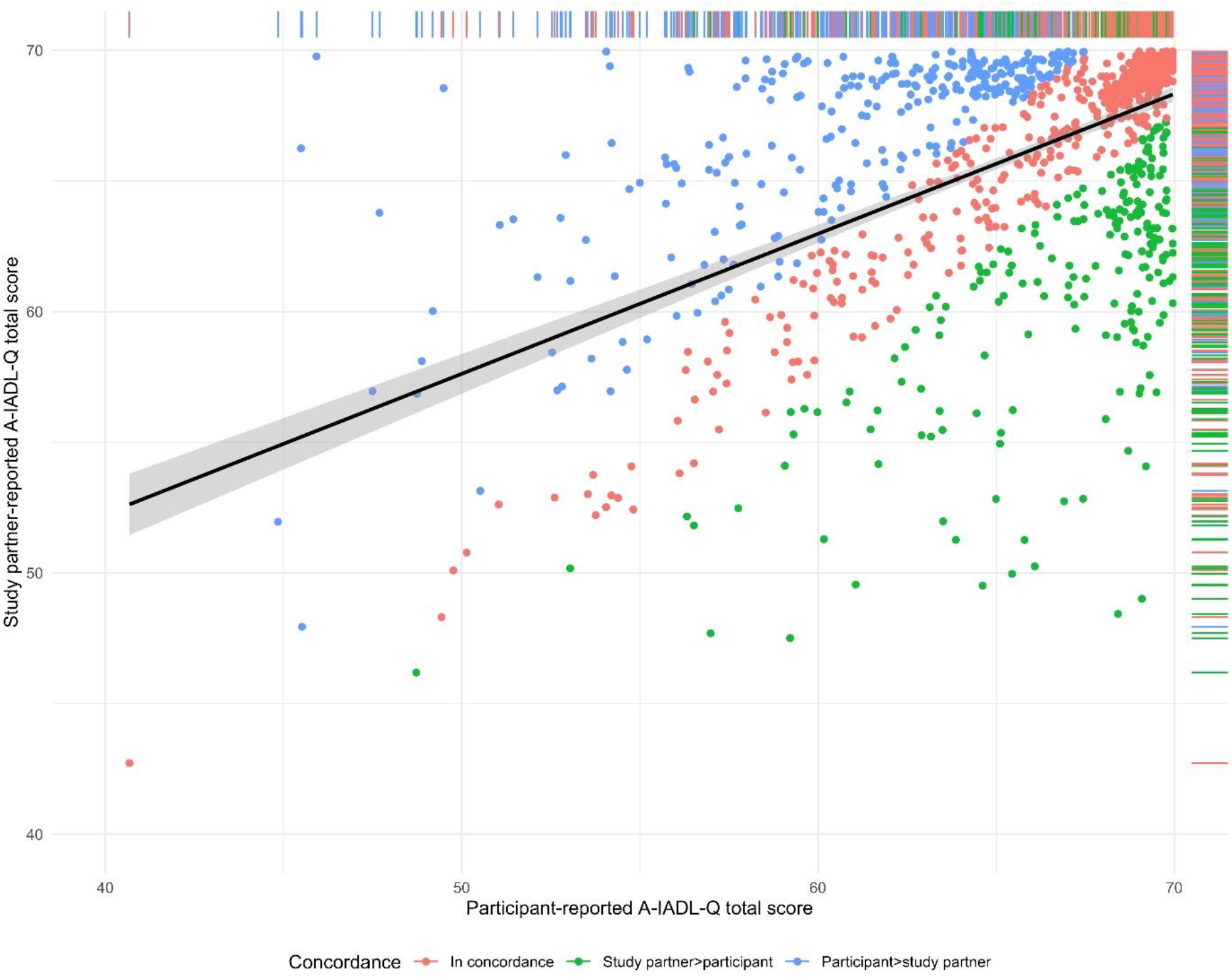
Scatterplot showing the relationship (black line) between participant-reported (horizontal axis) and study partner-reported IADL functioning (vertical axis). Each dot represents an individual; dots are colored based on a difference in IADL-Q scores of 2.4 points or more: dyads in concordance are red, dyads where the study partner reported better A-IADL-Q scores than the participant are green, dyads where the participant reported better A-IADL-Q scores than the study partner are blue.

